# A qualitative phase I study protocol on developing patient-reported outcome measure for hair, scalp, eyebrows, eyelashes, and beard-related skin disorders

**DOI:** 10.1101/2022.08.14.22278626

**Authors:** Yu-Feng Chang, Lara Drake, Sophia Reyes-Hadsall, John Barbieri, Arash Mostaghimi

## Abstract

**Introduction:** Hair disorders can have both physical and psychosocial effects significantly impacting patients’ quality of life. Well-designed patient-reported outcome measures (PROMs) are vital to determine meaningful patient-centered management. Currently available PROMs for individual hair disorders are available, however, making it difficult to use across diseases. This phase I protocol aims to create a clinically meaningful hair assessment across multiple hair disorders that evaluate the patients’ reported assessment of their disease, its symptoms, and impact on their quality of life, as well as defining characteristics of their desired outcome.

**Methods and analysis:** This phase I study, we will use thematic analysis of qualitative semi-structured interviews. Participants aged 18 years and older, a sampling of patients diagnosed with alopecia areata, androgenic alopecia, lichen planopilaris, central centrifugal cicatricial alopecia, and sebopsoriasis will be invited to take part in qualitative interviews. Participants recruitment and interviews will continue iteratively until thematic saturation is reached, with a focus on interview quality and patient diversity in gender, race and ethnicity, age, and hair-related disorder. The open-ended interview questions will allow participants to reflect on their physical appearance, disease journey, and treatment goals, as well as define the associated physical symptoms and psychosocial impact. These predominant domains will be used to form the basis for a hair-related PROM that can be applied to a multitude of hair-related disorders.

**Ethics and dissemination:** The Brigham and Women’s Hospital Institutional Review Board (2022P001033) approved this study. Following the description of the study protocol, we obtained verbal consent from all the study participants. All personal information, interview responses, and medical records are confidential and are stored in an encrypted database. Findings from this study will be published in peer-reviewed journals and presented at dermatology conferences.

This is an open access article with Creative Commons Attribution Non Commercial (CC BY-NC 4.0) license, that permits others to distribute, remix, and build upon the licensed work non-commercially, and require attribution to the original authors. See https://creativecommons.org/licenses/by-nc/4.0/ for details.

**Strengths and limitations of this study:**

- Recruitment of participants with common scarring and non-scarring hair-related disorders will make it possible to establish patients’ perspectives on physical appearance, definitions of treatment success, physical signs and symptoms, psychological and social impact across multiple disease states involving hair, scalp, eyebrows, eyelashes, and beard.
- Inclusion of patients from a wide range of demographics such as age, gender, race, and ethnicity will enable identification of domains common across these differing backgrounds.
- We adhere to published guidelines for determining validity of scales.
- We aim to use modern psychometric approach to ensure this study are clinically meaningful.

## Introduction

Hair disorders are complex, multi-faceted conditions that affect individuals both physically and psychologically. Due to their relapsing-remitting clinical course, unpredictable outcomes, and often long treatment courses, hair disorders significantly affect individuals’ psychological and social well-being and have been linked with psychiatric comorbidities like anxiety and depression.^1^ As shared decision-making is essential for navigating the evolving treatment landscape in hair disorders, clarification of patients’ perspectives and unique needs are crucial in the development of novel treatment modalities, especially regarding impacts on quality of life.

Patient-reported outcome measures (PROMs) are a valuable tool to capture the patient’s voice and to understand their lived experience of hair disorders. In clinical trials, PROMs are vital to determine if the study intervention is meeting the needs and priorities of the patients, which may differ from Clinician-Reported Outcome Measures (ClinROs).^2^ In clinical practice, PROMs are an ideal tool to evaluate the lived experience and unique treatment goals as part of shared decision making.

An ideal PROM will incorporate the patient’s assessment of their disease, symptoms, and impact on their quality of life.^3^ There are PROMs for a variety of specific hair disorders such as alopecia areata (AA),^4,5^ psoriasis,^6^ and scalp dermatitis,^7^ among others. However, there is currently no standardized universal PROM that encompasses all hair conditions. The lack of standardization and multitude of PROMs employed in clinical studies makes it difficult to compare treatment outcomes across trials and hair conditions. In addition, many of these PROMs were developed without advanced psychometric techniques such as Rasch Measurement Theory (RMT) and Item Response Theory, which have advantages over Classical Test Theory, including the ability to use computer adaptive testing to reduce response burden. In this study, we aim to create a clinically meaningful PROM with strong evidence for content validity and measurement properties using RMT.^8^

In this protocol, we describe the methodology to create an interview instrument and establish a protocol for the Phase I qualitative study. The interview guide and qualitative study protocol consider patient’s perceptions across different age, gender, races/ethnicities, and cultural backgrounds diagnosed with their disease will factor into the development of a clinically meaningful PROM assessment for use across all hair disorders.

## Methods and analysis

### Study design

To develop the interview instrument, a systematic review of the literature was performed to identify existing PROMs for hair-related disorders. This search was used to guide a preliminary conceptual framework that was then formulated into a preliminary structured interview guide. This work will be conducted in three phases. Phase I will involve generating a pool of domains identified using qualitative methods and developing these items into scales to be further refined by patients and experts. This phase ensures that the PROM will comprehensively encompass the patient-driven outcomes and have strong content validity.^11^ The initial interview guide was then pilot testing among patients with a hair disorder and assessed by both patients and experts for clarity incorporating patients’ feedback. The resulting qualitative interview guide will be administered in a series of semi-structured, open-ended interviews to a purposefully sampled group of individuals with hair disorders encompassing a diverse range of demographics and hair disorders. Phases II and III will involve measurement of the quantitative domains identified using field testing and assessing the measurement properties of the scales using psychometric analysis, respectively.

Qualitative data will be used to generate a set of domains or subthemes that characterize patients’ perceptions on values, satisfactory hair-related outcomes, and the physical and psychosocial effects of hair disorders, thereby completing Phase I. We followed guidance from the US Food and Drug Administration (FDA) on developing PROMS^12^ and from COnsensus-based Standards for the selection of health Measurements INstruments (COSMIN).^13^ We will follow the Consolidated Criteria for Reporting Qualitative Research (COREQ) reporting guideline for qualitative studies.^14^ Data collected through interviews with the patients will be analyzed with Interpretive Description (ID), approach used to generate knowledge relevant to the clinical context informing a study.^15^ We will conduct a series of semi-structured, open-ended interviews with a reference interview guide (Box 1) adapting to studies from Aldhouse et al^16^ and revising items as needed to identify concepts for scale development.

### Sample

Enrollment will be based on a purposefully sampled group of individuals of different age, gender, races/ethnicity, and education level diagnosed with the following diagnosis: alopecia areata (AA), androgenic alopecia (AGA), lichen planopilaris (LPP), central centrifugal cicatricial alopecia (CCCA), or sebopsoriasis, experiencing hair, scalp, eyelashes, eyebrows, beard-related dermatological conditions. Verbal consent will be obtained, and participants will be compensated for their time and effort. Participants will be recruited from the Brigham and Women’s Hospital, Department of Dermatology. Each individual interview will be conducted either in-person or through Zoom will be led by an experienced and trained member of the study team members.

### Part I: Development of Qualitative Interview Instrument

Development of this study followed a systematic methodologic review by Kallio et al^17^, the 5-step process presented in Figure. 1. First, evaluate appropriateness of the interview, determining the content is meaningful and focusing on patients’ values. Second, a literature review was conducted to identify existing PROMs for hair/scalp, eyebrows, eyelashes, and beard-related hair conditions. Based on expert input and a systematic review of existing PROMs for hair/scalp, eyebrows, eyelashes, and beard-related hair conditions, a semi-structured interview guide was developed. This interview guide was then pilot tested among patients with hair disorders and refined based on this feedback as well as input from an expert in qualitative research methodology. The final interview guides were then pilot testing once more for any additional feedback, Table 1.

**Figure 1.**
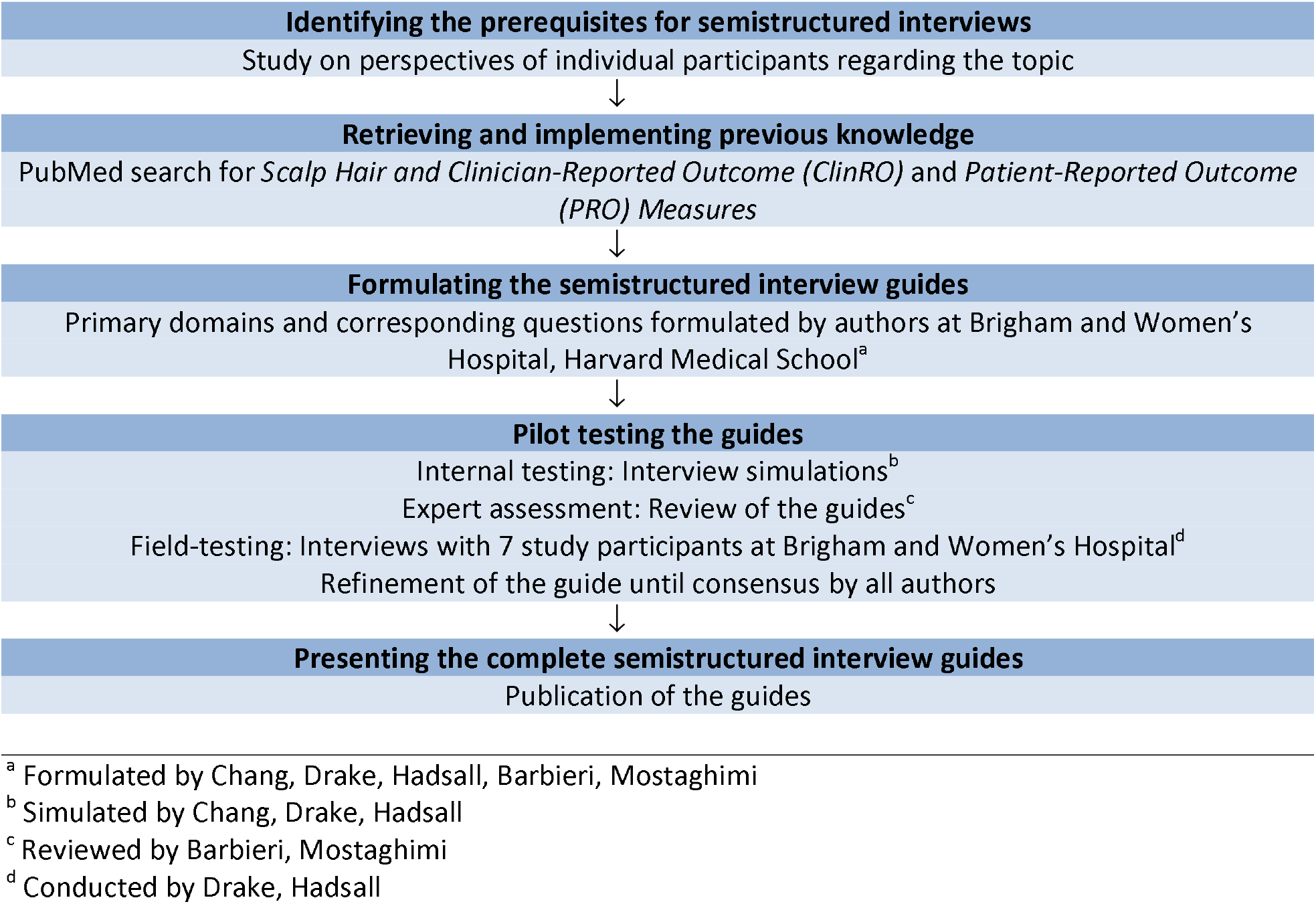
Flow Diagram for the Development of Semistructured Interview Guides.

**Table 1.** Framework of the study.

| Phase I | Purpose | Component | Product |
| --- | --- | --- | --- |
| Part I | What is lacking?<br>What to measure? | Systematic review<br>Interpretive description<br>Clinician input<br>Qualitative interviews to develop PROM | Preliminary framework<br>Refined conceptual framework<br>Preliminary scales |
| Part II | Measure the domains identified in Phase I | Pilot field testing | Refined scales |

### Part II: Protocol for Conducting Qualitative Interviews

Once the qualitative interview guide was finalized, a protocol for conducting the qualitative interviews was established. Purposeful sampling will be employed to ensure inclusion of patients of different gender, race/ethnicity, diagnosed with scarring (LPP, CCCA) and non-scarring (AGA, AA, sebopsoriasis) types of hair loss. This will ensure a diverse population representative of the scope of use for the PROM. Each patient will be introduced to the study by a dermatologist, after which additional information will be provided by study staff. Verbal consent will be obtained prior to recording the interview. All individuals will be interviewed with the same set of questions on their perspective on physical appearance, signs and symptoms, psychological and social impact related to their hair, scalp, eyebrows, eyelashes, and/or beard-related conditions. To minimize bias: 1) participants of different race/ethnicity, age, and gender will be recruited, 2) open-ended questions are framed carefully and reviewed by experts in the field for second opinion, and 3) data analysis and reporting will be supervised by an expert to ensure the good content validity and reliability.

We will also collect standardized information including the patient’s age, gender, race, ethnicity, and diagnosis from the patient. This method of data collection seeks to explore the patient’s views and perspectives, and our analysis will aim to define key concepts and points of consensus gathered during the conversation. Participants will be asked to answer open-ended questions on their own perspectives, using the guided questionnaire. Interviews will be performed by trained interviewers and transcribed verbatim.

### Sample size

The sample size for this study depends on the complexity of the PROM and the diversity of the sample population. Adequate sample size using group-based trajectory modeling depends on the dataset’s representativeness of the population of interest.^17^ We anticipate conducting a total of 12 interviews or until thematic saturation is reached. This open-ended, semi-structured interview uses the “think aloud method” with probing to obtain any missing content in each scale.^18^ This valuable approach ensure that respondents understood the content and tailor the scale accordingly.

### Statistical Analysis

The interview audio-recordings will be transcribed verbatim. Transcripts will be coded by two members of the research team per interview using a qualitative data management software system (NVIVO-11) (QSR International, Melbourne, Australia). Analysis will be mainly descriptive. Interviews (data collection) and coding (data analysis) will happen simultaneously to build on the domains identified from each interview. Thematic analysis will be used to code the domains that reoccur or appear important. For new concepts, codes will assign top-level domains and themes/subthemes to participant quotes. The resulting themes and subthemes identified from these qualitative interviews will be used to refine the conceptual framework, and to develop an item pool to inform preliminary scale development. This study will use a grounded theory approach to develop the code book. Authors will independently code the first 12 interviews, and codes will be refined until consensus is reached. Team members will independently finalize the codes, and all discrepancies will be discussed among all team members. Continuous variables will be summarized with means and standard deviations (SDs). Categorical variables are reported as proportions and percentages. Interrater reliability was assessed using the Cohen K coefficient. Qualitative and quantitative analyses will be performed in NVivo, version 12.1 (QSR International).

### Patient and public involvement

Prior to launching the study, the interview guide was pilot tested with 7 patients diagnosed with AA (4), LPP (1), and sebopsoriasis (2) to evaluate whether the PROM instrument would capture their experience and perception. All patients agreed that the frequency of interview questions and the length of the interview questions were reasonable and important. All participants in the initial qualitative interviews will be invited to continue to collaborate in our study during the refinement interviews. For all participants, we will provide feedback in form of a newsletter with information regarding the presentations and publications of the study.

### Ethics

Each participant will provide verbal consent before participating, participants will be compensated for their time and effort. Given that hair-related disorders including AGA, AA, LPP, CCCA, and sebopsoriasis are chronic, difficult to treat, and known to adversely affect patients’ quality of life, these interview questions may lead to emotional and psychological discomfort for the participants when talking about their experience. Participants can choose to discontinue the interview or skip questions. Patient data will be de-identified, secured, and confidential following the Brigham and Women’s Hospital Institutional Review Board rules for data storage.

### Dissemination

To ensure that this hair-related PROM can be universally used, once this instrument is developed, our team will present this study at dermatological conferences in both national and international meetings. We will promote its use among stakeholders including patients, physicians, and clinical trial investigators. Our dissemination initiatives will include in-person interactions and electronic media, all effective strategies for developing a well-represented PROM. We will publish our findings in peer-reviewed journals to raise awareness and promotes the use of our work

#### Box 1. Semi-structured interview guide

**Hair/Scalp**

A. **Appearance**
  1. Think about a time, either before your [scalp/hair condition] or more recently when your hair/scalp was at its best. What <u>qualities/specific aspects of your hair</u> made you feel that way?
    - Probe: appearance/texture/color, made it at its best?
  2. Describe how your hair has changed compared to <u>before</u> your [hair/scalp disease] or [if you cannot think that far back] compared to when it was recently at its <u>best</u>.
  3. What does treatment success look like to you?
    - Probe: What are your treatment goals?
B. **Feel/Symptoms**
  1. Think about a time when your [hair/scalp condition] symptoms were at their worst. How did your scalp <u>feel</u>?
    - Probe: pain: burning/stinging, itch
    - Probe: redness, scaling/flaking, cracking, bleeding
  2. At its worst, how would you describe the severity of your symptoms?
    - Probe: mild, moderate, severe, or scale [if severe/8-10, why?] [if moderate/4-7, why not severe/mild?] [if mild/0-3, why not severe?]
  3. At its worst, how frequent were your symptoms?
    - Probe: not at all, sometimes, most of the time, all the time
C. **Psychological**
  1. Think about your [hair/scalp condition] at its <u>worst:</u>
    1. Can you describe how you felt about yourself?
      - Probe: depressed, frustrated, anxious, ashamed/embarrassed
    2. At its worst, can you describe how your [hair/scalp condition] impacted:
      - Your self-confidence or self-image?
      - How did it impact the way you thought others perceived you or how you presented yourself to others?
      - How did it affect your perceived attractiveness?
  2. Think about either prior to your [hair/scalp condition] or when it was at its <u>best</u>:
    - Can you describe how you felt about yourself?
    - Can you describe how your self-confidence or self-image changed?
    - Think about when your [hair/scalp condition] was at its <u>best</u>. How did it change the way you thought others perceived you or how you presented yourself to others?
    - How did it affect your perceived attractiveness?
  3. How would failing [how did failing] a treatment make you feel?
  4. [If achieved remission]: how would you feel if your [hair/scalp condition] returned
D. **Social**
  1. When your [hair/scalp condition] was at its <u>worst</u>:
    1. What actions or behaviors did you implement to make yourself feel/look better or to conceal your condition?
      - Probe: Accessories, hats, wigs, haircuts, hairstyles, dyeing
      - Probe: How much effort have you put into your hair due to your [hair/scalp condition]?
  2. Can you describe any changes you had to make to your daily life when your [hair/scalp condition] was at its <u>worst</u>?
    1. [Work/School]: How did this impact your contributions in a work/school environment?
    2. [Daily/Leisure activities/social life]:
      1. How did it impact your social life?
      2. How did it impact your leisure activities?
        - Probe: Did you have to change any of your preferred leisure activities?
        - Probe: Did it impact the time spent doing these leisure activities?

**Eyebrow**

A. **Appearance**
  1. Think about a time, either before your [eyebrow condition] or more recently when your eyebrows were at their best. What <u>qualities/what specific aspects of your eyebrows</u> made you feel that way?
    - Probe: color/symmetry/length/shape
  2. Describe how your eyebrows have changed compared to <u>before</u> your [eyebrow condition] or [if you cannot think that far back] compared to when it was recently at their <u>best</u>.
  3. Describe how your ideal eyebrows look after treatment.
    - Probe: What are your treatment goals?
B. **Feel/Symptoms**
  1. Think about a time when your [eyebrow condition] symptoms were at their worst. How did your eyebrows <u>feel</u>?
    - Probe: pain, itch, bleeding
  2. At its worst, how would you describe the severity of your symptoms?
    - Probe: mild, moderate, severe or scale [if severe/8-10, why?] [if moderate/4-7, why not severe/mild?] [if mild/0-3, why not severe?]
  3. At its worst, how frequent were your symptoms?
    - Probe: not at all, sometimes, most of the time, all the time
C. **Psychological**
  1. Think about your [eyebrow condition] at its <u>worst:</u>
    1. Can you describe how you felt about yourself?
      - Probe: depressed, frustrated, anxious, ashamed/embarrassed
    2. At its worst, can you describe how your [eyebrow condition] impacted:
      - Your self-confidence or self-image?
      - How did it impact the way you thought others perceived you or how you presented yourself to others?
      - How did it affect your perceived attractiveness?
  2. Think about either prior to your [eyebrow condition] or when it was at its <u>best</u>:
    1. Can you describe how you felt about yourself?
    2. Can you describe how your self-confidence or self-image changed?
  3. Think about when your [eyebrow condition] was at its <u>best</u>. How did it change the way you thought others perceived you or how you presented yourself to others?
    - How did it affect your perceived attractiveness?
  4. How would failing [how did failing] a treatment make you feel?
  5. [If achieved remission]: how would you feel if your [eyebrow condition] returned
D. **Social**
  1. When your [eyebrow condition] was at its <u>worst</u>:
    1. What actions or behaviors did you implement to make yourself feel/look better or to conceal your condition?
      - Probe: Haircut (i.e., bangs), tattooing, makeup, tinting/dyeing
      - Probe: How much effort have you put into your eyebrows due to your [eyebrow condition]?
  2. Can you describe any changes you had to make to your daily life [work/school/leisure activities] when your [eyebrow condition] was at its <u>worst</u>?
    1. [Work/School]: How did this impact your contributions in a work/school environment?
    2. [Daily/Leisure activities/social life]:
      1. How did it impact your social life?
      2. How did it impact your leisure activities?
        - Probe: Did you have to change any of your preferred leisure activities?
        - Probe: Did it impact the time spent doing these leisure activities?

**Eyelashes**

A. **Appearance**
  1. Think about a time, either before your [eyelash condition] or more recently when your eyelashes were at their best. What <u>qualities/specific aspects of your eyelashes</u> made you feel that way?
    - Probe: length/thickness/color
  2. Describe how your eyebrows have changed compared to <u>before</u> your [eyelash condition] or [if you cannot think that far back] compared to when it was recently at their <u>best</u>.
  3. Describe how your ideal eyelashes look after treatment.
    - Probe: What are your treatment goals?
B. **Feel/Symptoms**
  1. Think about a time when your [eyelash condition] symptoms were at their worst. How did your eyelashes <u>feel</u>?
    - Probe: pain, itch, bleeding
  2. At its worst, how would you describe the severity of your symptoms?
    - Probe: mild, moderate, severe or scale [if severe/8-10, why?] [if moderate/4-7, why not severe/mild?] [if mild/0-3, why not severe?]
  3. At its worst, how frequent were your symptoms?
    - Probe: not at all, sometimes, most of the time, all the time
C. **Psychological**
  1. Think about your [eyelash condition] at its <u>worst:</u>
    1. Can you describe how you felt about yourself?
      - Probe: depressed, frustrated, anxious, ashamed/embarrassed
    2. At its worst, can you describe how your [eyelash condition] impacted:
      - Your self-confidence or self-image?
      - How did it impact the way you thought others perceived you or how you presented yourself to others?
      - How did it affect your perceived attractiveness?
  2. Think about either prior to your [eyelash condition] or when it was at its <u>best</u>:
    1. Can you describe how you felt about yourself?
    2. Can you describe how your self-confidence or self-image changed?
  3. Think about when your [eyelash condition] was at its <u>best</u>. How did it change the way you thought others perceived you or how you presented yourself to others?
    - How did it affect your perceived attractiveness?
  4. How would failing [how did failing] a treatment make you feel?
    1. [If achieved remission]: how would you feel if your [eyelash condition] returned
D. **Social**
  1. When your [eyebrow condition] was at its <u>worst</u>:
  2. What actions or behaviors did you implement to make yourself feel/look better or to conceal your condition?
    - Probe: Haircut (i.e., bangs), tattooing, makeup, tinting/dyeing
    - Probe: How much effort have you put into your eyebrows due to your [eyebrow condition]?
  3. Can you describe any changes you had to make to your daily life [work/school/leisure activities] when your [eyelash condition] was at its <u>worst</u>?
    1. [Work/School]: How did this impact your contributions in a work/school environment?
    2. [Daily/Leisure activities/social life]:
      1. How did it impact your social life?
      2. How did it impact your leisure activities?
        - Probe: Did you have to change any of your preferred leisure activities?
        - Probe: Did it impact the time spent doing these leisure activities?

**Beard**

A. **Screening question:** Do you have experience with having a beard or wanting to grow a beard?
B. **Appearance**
  1. Think about a time, either before your [beard condition] or more recently when your beard was at its best. What <u>qualities/</u>specific aspects of your beard made you feel that way?
    - Probe: color/length/thickness/shape
  2. Describe how your beard has changed compared to <u>before</u> your [beard condition] or [if you cannot think that far back] compared to when it was recently at its <u>best</u>.
  3. Describe how your ideal beard looks after treatment.
    - Probe: What are your treatment goals?
C. **Feel/Symptoms**
  1. Think about a time when your [beard condition] symptoms were at their worst. How did your beard <u>feel</u>?
    - Probe: pain: burning/stinging, itch
    - Probe: redness, scaling/flaking, cracking, bleeding
  2. At its worst, how would you describe the severity of your symptoms?
  3. Probe: mild, moderate, severe or scale [if severe/8-10, why?] [if moderate/4-7, why not severe/mild?] [if mild/0-3, why not severe?]
  4. At its worst, how frequent were your symptoms?
    - Probe: not at all, sometimes, most of the time, all the time
D. **Psychological**
  1. Think about your [beard condition] at its <u>worst:</u>
    1. Can you describe how you felt about yourself?
      - Probe: depressed, frustrated, anxious, ashamed/embarrassed
  2. At its worst, can you describe how your [beard condition] impacted:
    1. Your self-confidence or self-image?
    2. How did it impact the way you thought others perceived you or how you presented yourself to others?
      i. How did it affect your perceived attractiveness?
    3. Think about either prior to your [beard condition] or when it was at its <u>best</u>:
      i. Can you describe how you felt about yourself?
      ii. Can you describe how your self-confidence or self-image changed?
    4. Think about when your [beard condition] was at its <u>best</u>. How did it change the way you thought others perceived you or how you presented yourself to others?
      i. How did it affect your perceived attractiveness?
    5. How would failing [how did failing] a treatment make you feel?
    6. [If achieved remission]: how would you feel if your [beard condition] returned
E. **Social**
  1. When your [beard condition] was at its <u>worst</u>:
    1. What actions or behaviors did you implement to make yourself feel/look better or to conceal your condition?
      - Probe: Shaving, hair growth products, dyeing
      - Probe: How much effort have you put into your beard due to your [beard condition]?
  2. Can you describe any changes you had to make to your daily life when your [beard condition] was at its <u>worst</u>?
  3. Can you describe any changes you had to make to your daily life [work/school/leisure activities] when your [beard condition] was at its <u>worst</u>?
    1. [Work/School]: How did this impact your contributions in a work/school environment?
    2. [Daily/Leisure activities/social life]:
      1. How did it impact your social life?
      2. How did it impact your leisure activities?
        - Probe: Did you have to change any of your preferred leisure activities?
        - Probe: Did it impact the time spent doing these leisure activities?

## Data Availability

All data produced in the present work are contained in the manuscript.

